# Hormone replacement therapy and dementia risk among postmenopausal women: evidence from the UK Biobank

**DOI:** 10.1101/2025.07.22.25331871

**Authors:** Steven Squires, Rasha NM Saleh, Luke C Pilling, Janice L Atkins, Janice M Ranson, Xin You Tai, David Vauzour, David J Llewellyn, Anne-Marie Minihane

## Abstract

**Importance:** The relationship between HRT use and dementia risk is unclear, with studies reporting both increased and decreased risk.

**Objective:** To identify modulators of the association between HRT use and dementia risk and identify potentially responsive subgroups of women.

**Design, setting, and participants:** This study used data from the UK Biobank, with baseline assessment conducted between 2006 and 2010 and follow up through to October 2022. The cohort included post-menopausal women without a diagnosis of dementia at baseline.

**Exposures:** The primary exposure was HRT use for a minimum duration of one year.

**Main outcomes and measures:** Multivariable Cox proportional hazards regression models were employed to evaluate the relationship between HRT use and incident all-cause dementia, Alzheimer’s disease (AD) and non-AD dementia. Stratified analyses were conducted to explore heterogeneity in the associations and the impact of potential effect modifiers, including type of menopause (surgical vs. natural), cumulative lifetime exposure to natural estrogen and *APOE ε4* genotype. The association between age at HRT initiation and dementia risk was also examined.

**Results:** Among 183,450 postmenopausal women (mean [SD] age at baseline, 60.3 [5.8] years), there were 3,948 dementia cases over 2,433,320 person-years of follow-up. Of these, 1,993 (50.5%) were classified as AD and 1,955 (49.5%) as non-AD dementia. HRT use was associated with lower risk of all-cause dementia (hazard ratio [HR], 0.90; 95% CI, 0.84-0.96). Stronger associations were observed in women with surgical menopause (HR 0.76; 95% CI, 0.67-0.85), *APOE ε4* carriers (HR 0.87; 95% CI, 0.80-0.95), and those with a lower lifetime exposure to natural estrogen (HR 0.78; 95% CI, 0.70-0.86). Initiation of HRT between 46 and 56 years was associated with reduced dementia risk.

**Conclusions and relevance:** In this large UK Biobank cohort, HRT use was associated with reduced risk of dementia among post-menopausal women. The association varied by age of initiation of HRT, *APOE* genotype, type of menopause, and endogenous lifetime estrogen exposure, highlighting the importance of personalised approaches to HRT prescribing. While further research is required to establish causality, the findings offer a foundation for tailored interventions aimed at mitigating lifetime dementia risk in women.

**Key Points:** *Question:* Is hormone replacement therapy (HRT) use associated with dementia risk in women?

*Findings:* In this prospective cohort study involving 183 450 post-menopausal women, HRT use for one year or more was associated with a reduced dementia risk. The risk reductions were more pronounced for women who had surgical menopause, had a shorter lifetime exposure to natural estrogen, or have at least one *APOE* ε4 allele. Notably the protective associations were most evident in those who initiated HRT between 46 and 56 years.

*Meaning:* HRT use was associated with an overall reduction in dementia risk. The likelihood of benefit is nuanced and dependant on several common variables, highlighting the importance of a more personalised approach to HRT prescribing.

## Introduction

Age-standardised dementia rates are higher in women compared to men, suggesting that female-specific risk factors may contribute to this disparity^1^. In premenopausal women, estrogen plays a critical role in supporting brain function through its effects on synaptic integrity, neurotransmitter metabolism, glucose utilisation, blood brain barrier maintenance, cerebrovascular function and neuroinflammation^2,3^. The menopausal transition represents an endocrine inflection point, that often coincides with the onset of the preclinical phase of dementia^4^, and surgically induced menopause via oophorectomy has been linked with an increased life-time risk of dementia.^5,6^

Despite biological plausibility, evidence regarding the association between hormone replacement therapy (HRT), which is used by approximately 15% of women aged 45-64y in England, and cognitive outcomes or dementia risk is conflicting and inconclusive^7,8^ as highlighted in the most recent meta-analysis.^9^ The only RCT to date assessing incident dementia as a primary outcome, the Women’s Health Initiative Memory Study (WHIMS), reported an increased risk of dementia with HRT use^10^, with several other studies concluding similarly^11–14^. Conversely, HRT has also been found to be both protective^15–17^ and to have no effect^18,19^.

These discrepancies may reflect the influence of multiple modifying factors, many of which remain poorly characterised. The APOE ε4 allele, the most significant common genetic risk factor for dementia^20–22^, exhibits higher penetrance in women^23^, and may influence the cognitive effects of HRT^24,25^. Associations between HRT use and dementia could also differ by age at menarche and menopause as these alter the lifetime estrogen exposure in women with evidence that fewer years of exposure is associated with greater neuropathological burden^26^ and dementia risk^18,27,28 29^. Surgical menopause may further increase risk and alter responsiveness to HRT^5,30^. Moreover, the “critical window” hypothesis proposes that HRT may be neuroprotective when initiated in the perimenopausal or early postmenopausal period^31,32^ though this has not been systematically investigated.

The aim of the present study was to use data from a large population-based cohort to identify potential effect modifiers and subgroups that may derive benefit or harm from HRT use.

## Methods

### Study design and participants

We used data from UK Biobank, an ongoing cohort study of more than 500,000 people aged 37 to 73 years at baseline^33^. Participants were assessed at 22 assessment centres around the United Kingdom between 2006 and 2010.

Analyses were restricted to post-menopausal women who experienced either natural (spontaneous) or surgical menopause (hysterectomy and/or bilateral oophorectomy). Women who had dementia at baseline, were missing information on HRT (whether they ever used HRT or their age at first or last use) or menopausal status were excluded. We followed the Strengthening the Reporting of Observational Studies in Epidemiology (<u>STROBE</u>) guidelines.

### Incident dementia

Our primary outcome was date first diagnosed with ‘all-cause dementia’, with Alzheimer’s disease (AD) and non-AD dementia (defined as ‘all-cause dementia’ excluding participants who ever received an AD diagnostic code) as secondary outcomes. The reference group are participants with no dementia diagnosis. The censoring date for time to event analyses was set to the 30 October 2022 or date of death, whichever was earlier. Details of ascertainment of dementia diagnoses are in the Supplement.

### HRT use

We defined use of HRT, from self-reported data, as a binary variable, with current users of HRT, and those who had used HRT for at least one year as the exposure categories, and those who never used HRT or used HRT for less than 12 months as the reference category.

### Covariates

We adjusted the model for: age; systolic blood pressure; body mass index (BMI); serum cholesterol levels; Townsend deprivation index^34^ (all continuous variables); education as five categories; ethnicity as six categories; smoking status categorised as never smoked or smoker; diabetes defined as a binary variable; use of cholesterol lowering medication; use of anti-hypertensive medication. A specification of the covariates is provided in eTable1 of the Supplement. In addition, an unadjusted model with no covariates and a partially adjusted model with age, education and smoking status as covariates were trained and results reported in the Supplement.

### Statistical analysis

We applied Cox proportional hazard models to estimate the hazard ratios for dementia risk. Multiple stratified models were utilised with HRT use as one category and non-HRT use as the reference. Confidence intervals are reported at the 95% level and p-values have been reported where appropriate. Cox models were run using Python package *lifelines*^*35*^.

We investigated possible modifying effects of the following covariates: APOE ε4 status, surgical menopause, and length of natural estrogen exposure (time between menarche and menopause). APOE ε4 status is available in the genetic data of UK Biobank via two single-nucleotide polymorphisms: rs429358 and rs7412. A binary variable was derived for APOE status: no ε4 alleles or 1-2 ε4 alleles. For women who had surgical menopause we defined the age at menopause as the earliest age of either surgery or stated age at cessation of periods. If women were missing data on age of menarche or age of menopause they were excluded from those sub-analyses.

To explore the window of opportunity hypothesis we estimated the hazard ratios for women starting HRT at different ages compared to the reference category. We considered all women who started HRT earlier/later than a specified age from 40 to 59 years; for example, when considering earlier starting ages of HRT, we took all women who started HRT at 44 years or younger and assessed their risk of dementia compared to women who never took HRT. In addition, we assessed dementia risk for those women who started HRT within time windows of 46-50 years and 51-56 years (inclusive).

## Results

From the original sample of 273,155 women, 89,705 were excluded as they had not undergone menopause or had missing information on menopause (76,453), missing information on HRT (13,154) or because they had dementia at baseline (98), leaving 183,450 included for analysis (see eFigure 1 in the Supplement). Baseline characteristics are shown in Table 1. During 2,433,320 (mean [SD] follow-up duration 13.26 [1.96]; median [IQR] follow-up of 13.58 [12.83-14.33]) person-years of follow-up, there were 3,948 dementia diagnoses, of those 1,993 were Alzheimer’s disease and 1,955 were either “dementia, unspecified” or a non-Alzheimer’s form of dementia (non-AD).

**Table 1.** Baseline Characteristics.

| Characteristic | Incident dementia (n=3 948) | No dementia (n=179 502) | Total (n=183 450) |
| --- | --- | --- | --- |
| Age, mean (SD), y | 65.0 (4.1) | 60.1 (5.8) | 60.3 (5.8) |
| APOE status |  |  |  |
| No E4 Allele, n (%) | 1 614 (40.9) | 125 664 (70.0) | 127 278 (69.4) |
| 1 or 2 E4 Alleles, n (%) | 2 167 (54.9) | 48 020 (26.8) | 50 187 (27.4) |
| Menopause status<sup>a,b</sup> |  |  |  |
| Natural menopause, n (%) | 2 782 (70.5) | 133 222 (74.2) | 136 004 (74.1) |
| Age at natural menopause, mean (SD), y | 50.0 (4.9) | 50.3 (4.4) | 50.3 (4.4) |
| Surgical menopause, n (%) | 1 166 (29.5) | 46 280 (25.8) | 47 446 (25.9) |
| Age at surgical menopause, mean (SD), y | 43.5 (7.7) | 43.2 (7.1) | 43.2 (7.1) |
| Natural estrogen exposure<sup>c</sup> |  |  |  |
| Natural estrogen exposure, mean (SD), y | 35.0 (6.8) | 35.5 (6.3) | 35.5 (6.3) |
| Longer natural estrogen >34 y, n (%) | 2 182 (55.3) | 109 859 (61.2) | 112 041 (61.1) |
| Shorter natural estrogen <35 y, n (%) | 1 416 (35.9) | 59 566 (33.2) | 60 982 (33.2) |
| HRT use<sup>d</sup> |  |  |  |
| No HRT use, n (%) | 2 177 (55.1) | 103 938 (57.9) | 106 115 (57.8) |
| Used HRT, n (%) | 1 771 (44.9) | 75 564 (42.1) | 77 335 (42.2) |
| Education<sup>e,f</sup> |  |  |  |
| Other, n (%) | 1 349 (34.2) | 36 057 (20.1) | 37 406 (20.4) |
| Lower secondary, n (%) | 1 480 (37.5) | 89 038 (49.6) | 90 518 (49.3) |
| Upper secondary, n (%) | 596 (15.1) | 41 522 (23.1) | 42 118 (23.0) |
| NVQ, n (%) | 354 (9.0) | 22 942 (12.8) | 23 296 (12.7) |
| Higher, n (%) | 1 326 (33.6) | 80 010 (44.6) | 81 336 (44.3) |
| Smoking status<sup>f</sup> |  |  |  |
| Never smoked, n (%) | 2 135 (54.1) | 103 864 (57.9) | 105 999 (57.8) |
| Current or past smoker, n (%) | 1 783 (45.2) | 75 012 (41.8) | 76 795 (41.9) |
| Ethnicity<sup>f</sup> |  |  |  |
| White, n (%) | 3 804 (96.4) | 171 821 (95.7) | 175 625 (95.7) |
| Asian, n (%) | 42 (1.1) | 2 537 (1.4) | 2 579 (1.4) |
| Black, n (%) | 58 (1.5) | 2 365 (1.3) | 2 423 (1.3) |
| Mixed, n (%) | 18 (0.5) | 854 (0.5) | 872 (0.5) |
| Chinese, n (%) | 4 (0.1) | 491 (0.3) | 495 (0.3) |
| Other, n (%) | 22 (0.6) | 1 434 (0.8) | 1456 (0.8) |
| Systolic blood pressure<sup>f</sup>, mean (SD) (mmHg) | 145.5 (20.0) | 140.3 (19.6) | 140.4 (19.7) |
| BMI<sup>f</sup>, mean (SD) (kg/m<sup>2</sup>) | 27.6 (5.3) | 27.2 (5.1) | 27.2 (5.1) |
| Diabetes<sup>f</sup> |  |  |  |
| No diabetes, n (%) | 3 540 (89.7) | 171 817 (95.7) | 175 357 (95.6) |
| Diabetes, n (%) | 398 (10.1) | 7 328 (4.1) | 7 726 (4.2) |
| Cholesterol<sup>f</sup> (SD) (mmol/l) | 6.0 (1.2) | 6.0 (1.1) | 6.0 (1.1) |
| Blood pressure medication<sup>f</sup> |  |  |  |
| No blood pressure medication, n (%) | 2 522 (63.9) | 140 099 (78.0) | 142 621 (77.7) |
| Blood pressure medication, n (%) | 1 335 (33.8) | 37 489 (20.9) | 38 824 (21.2) |
| Cholesterol drugs<sup>f</sup> |  |  |  |
| No cholesterol medication, n (%) | 2 668 (67.6) | 149 833 (83.5) | 152 501 (83.1) |
| Cholesterol medication, n (%) | 1 189 (30.1) | 27 755 (15.5) | 28 944 (15.8) |
| Townsend deprivation index<sup>f</sup>, mean (SD) | 27.6 (5.3) | 27.2 (5.1) | 27.2 (5.1) |

Abbreviations: APOE, apolipoprotein E; BMI, body mass index. <sup>a</sup>Surgical menopause is defined as having had a bilateral oophorectomy or hysterectomy. <sup>b</sup>Age at surgical menopause is the earliest of stated menopause age, age at hysterectomy or age at bilateral oophorectomy. <sup>c</sup>Natural estrogen exposure is years between menarche and earliest of natural or surgical menopause. <sup>d</sup>No HRT use is defined as never using HRT or using HRT but starting and stopping at the same age. <sup>e</sup>Used HRT is defined as using HRT for at least one year or still using HRT at baseline. <sup>f</sup>Educational categories are specified in the Supplement. Results sum to more than 100% because multiple qualifications could be specified. <sup>g</sup>Missing values were imputed using multiple imputations by chained equations with 40 imputations.

### Association between HRT and all-cause dementia

HRT was associated with a reduced risk of dementia with a hazard ratio (HR) of 0.90 (95% CI, 0.84-0.96, P=0.001) (Figure 1), in a Cox’s proportional hazards regression model adjusted for age, education, smoking status, systolic blood pressure, ethnicity, BMI, cholesterol, Townsend deprivation index, diabetes, and use of cholesterol lowering or anti-hypertensive medications. For women who experienced natural menopause, this association was not statistically significant with a HR of 0.94 (95% CI, 0.87-1.01, P=0.089). For women who underwent surgical menopause HRT was associated with a reduction in dementia risk with a HR of 0.76 (95% CI, 0.67-0.85, P<0.001). For women who were not *APOE ε4* carriers, the association of HRT with dementia was not significant while for those with at least one allele the HR was 0.87 (95% CI, 0.80-0.95, P=0.002). The association between HRT and dementia depended on the length of natural estrogen exposure; for women with more than 34 years between menarche and either natural menopause or surgical menopause, there was no association between HRT and dementia. Conversely, for women with fewer years of exposure to natural estrogen the HR was 0.78 (95% CI, 0.70-0.87, P<0.001).

**Figure 1.**
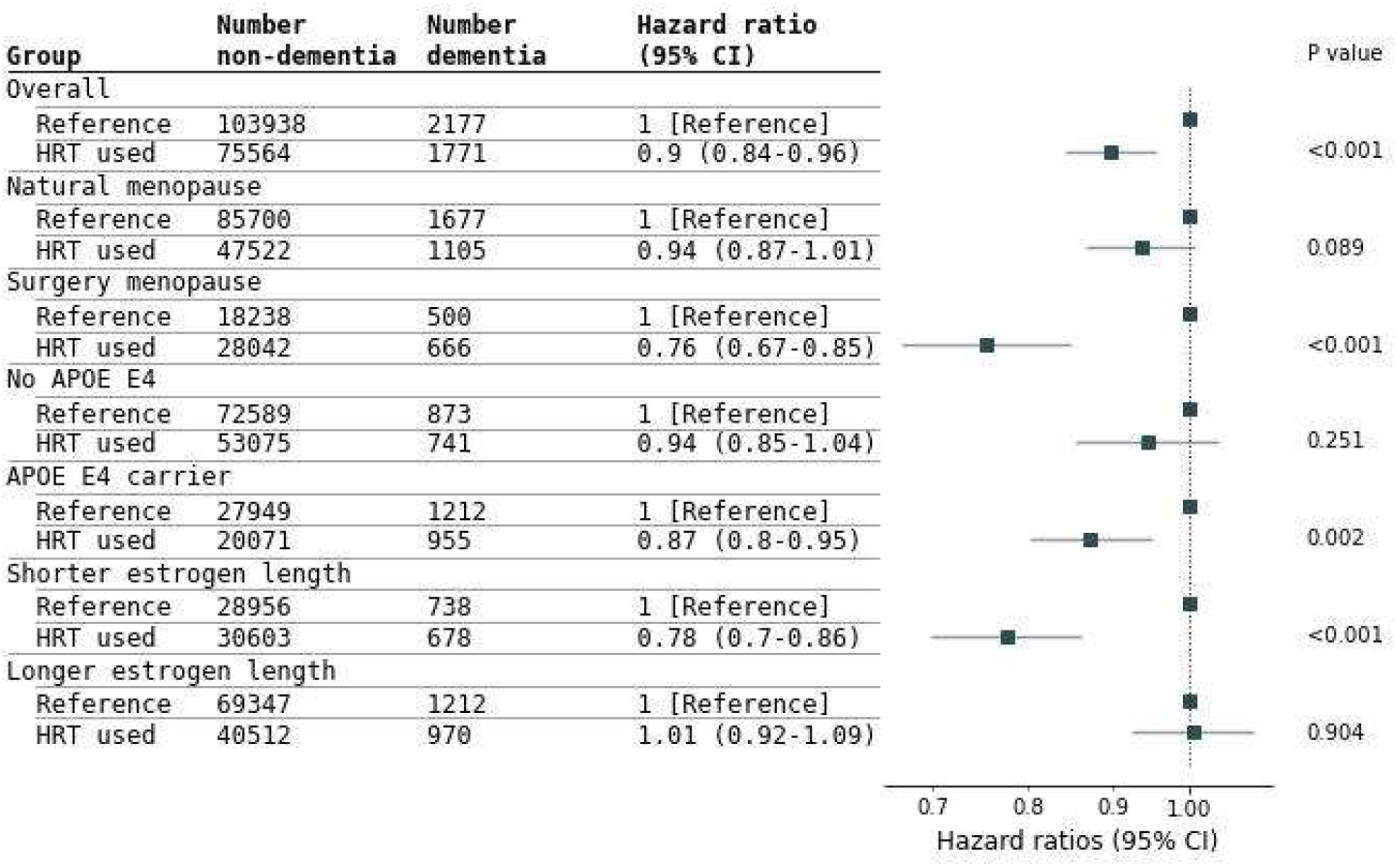
Risk of Incident Dementia Stratified by Groups. Abbreviations: APOE, apolipoprotein E. All results from a Cox proportional hazards regression adjusted for: age, education, smoking status, systolic blood pressure, ethnicity, BMI, cholesterol, Townsend deprivation index, diabetes, whether taking cholesterol lowering medication, whether taking anti-hypertensive medications. The reference for each group is those who never took HRT. The women defined as using HRT had used HRT for at least one year or were still taking HRT at baseline. Natural menopause is women who reported natural cessation of periods. Surgical menopause is women who had either a bilateral oophorectomy or hysterectomy. No APOE ε4 women did not have any ε4 alleles. APOE ε4 carriers had either one or two ε4 alleles. Estrogen length is defined as time between menarche and the earliest of age at menopause, age at hysterectomy or bilateral oophorectomy. Shorter estrogen length is less than 35 years of natural estrogen length. Longer estrogen length is 35 years or more of natural estrogen length.

### Combinations of menopause type, APOE ε4 status and length of exposure to natural estrogen

The association between HRT and dementia depends on the combined effects of menopause type (natural or surgical menopause), *APOE ε4* status and length of exposure to natural estrogen (eFigure2 and eFigure3 in the Supplement). For women who had a natural menopause, HRT is not associated with dementia risk if the woman had more than 34 years of exposure to natural estrogen with or without an *APOE ε4* allele. However, for those women who had a natural menopause, were *APOE ε4* carriers, and had a shortened exposure to natural estrogen, HRT was associated with reduced dementia risk with a HR of 0.79 (95% CI, 0.63-0.98, P=0.03).

For women who had surgical menopause the weakest association was 0.84 (95% CI, 0.60-1.20, P=0.33) for women with no *APOE ε4* alleles and a longer exposure to natural estrogen. The largest association was a HR of 0.68 (95% CI, 0.55-0.83, P<0.001) for women with at least one *APOE ε4* allele and a shorter exposure to natural estrogen.

### Association of HRT and AD

HRT was associated with reduced AD risk with a HR of 0.84 (95% CI, 0.77-0.92, P<0.001) (Figure 2). For women who had a natural menopause the HR was 0.89 (95% CI, 0.80-0.99, P=0.03) and for those who had surgical menopause the HR was 0.72 (95% CI, 0.61-0.85, P<0.001). The HR was similar for women with no *APOE ε4* allele or with at least one at 0.85 (95% CI, 0.73-1.00, P=0.05) and 0.84 (95% CI, 0.75-0.94, P=0.003) respectively. The HR for women exposed to fewer than 35 years of natural estrogen was 0.73 (95% CI, 0.62-0.84, P<0.001) while for those exposed to more than 34 years of natural estrogen the HR was 0.95 (95% CI, 0.84-1.07, P=0.40).

**Figure 2.**
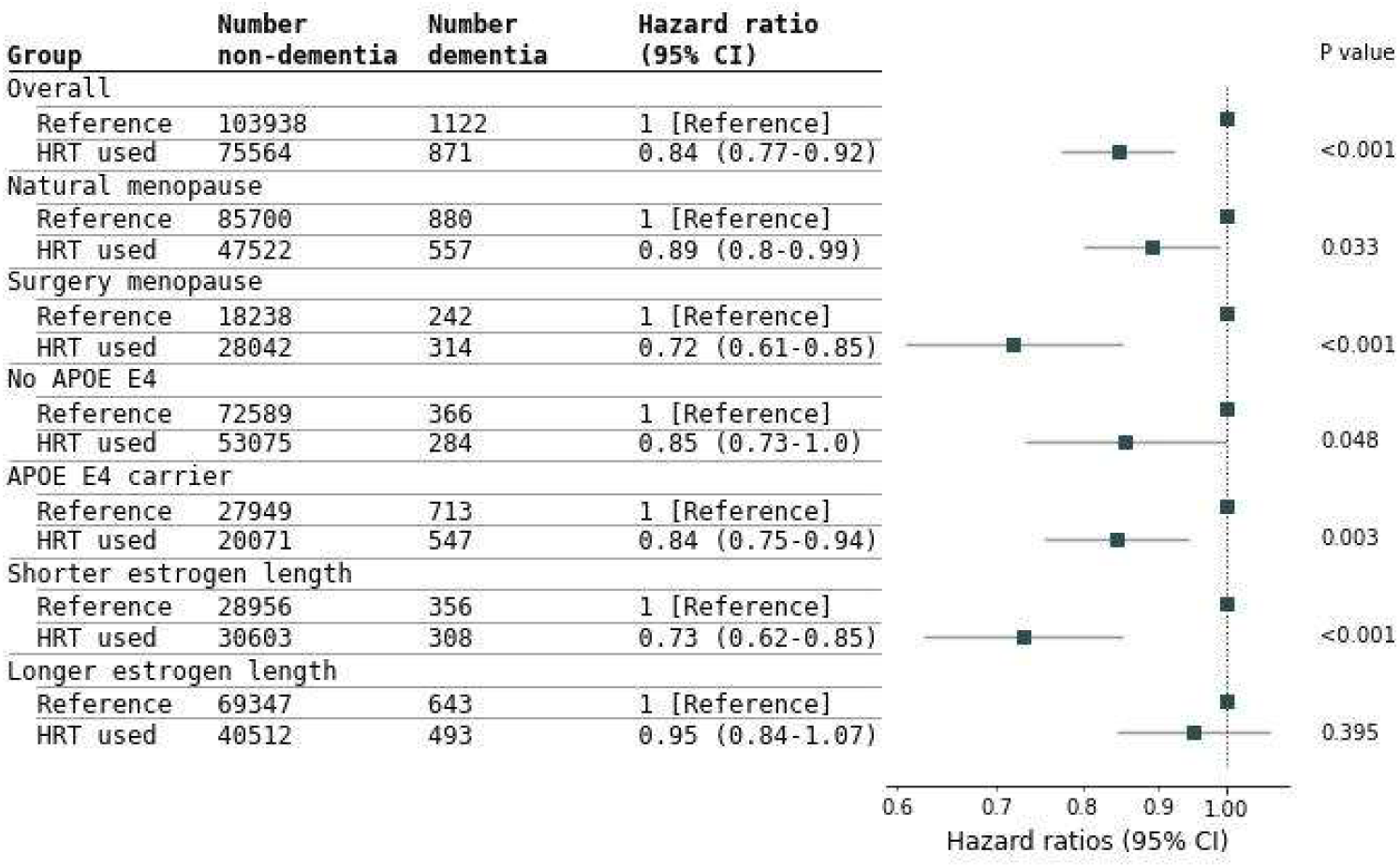
Risk of Incident AD Stratified by Groups. Abbreviations: APOE, apolipoprotein E. All results from a Cox proportional hazards regression adjusted for: age, education, smoking status, systolic blood pressure, ethnicity, BMI, cholesterol, Townsend deprivation index, diabetes, whether taking cholesterol lowering medication, whether taking anti-hypertensive medications. The reference for each group is those who never took HRT or took HRT but started and stopped at the same age. The HRT used women used HRT for at least one year or were still taking HRT at baseline. Natural menopause is women who reported cessation of periods and who had not have a bilateral oophorectomy or a hysterectomy. Surgical menopause are women who had either a bilateral oophorectomy or hysterectomy. No APOE ε4 women did not have any ε4 alleles. APOE ε4 carriers had either one or two ε4 alleles. Estrogen length is defined as time between menarche and the earliest of age at menopause, age at hysterectomy or age at bilateral oophorectomy. Shorter estrogen length is less than 35 years of estrogen length. Longer estrogen length is 35 years or more of estrogen length.

### Association of HRT and non-AD

There was no significant HRT effect for non-AD for all women, with a HR of 0.95 (95% CI, 0.87-1.04, P=0.28) (Figure 3) or for those who had a natural menopause with a HR of 0.99 (95% CI, 0.88-1.10, P=0.80). For those women who had surgical menopause the HR was 0.79 (95% CI, 0.67-0.93, P=0.004). The HRT association was not significant when stratifying into those with or without APOE ε4 alleles. For those women with fewer than 35 years of exposure to natural estrogen there was an association between HRT use and non-AD with a HR of 0.82 (95% CI, 0.71-0.94, P=0.006) while for those with more than 34 years of exposure to natural estrogen the HR was not significant.

**Figure 3.**
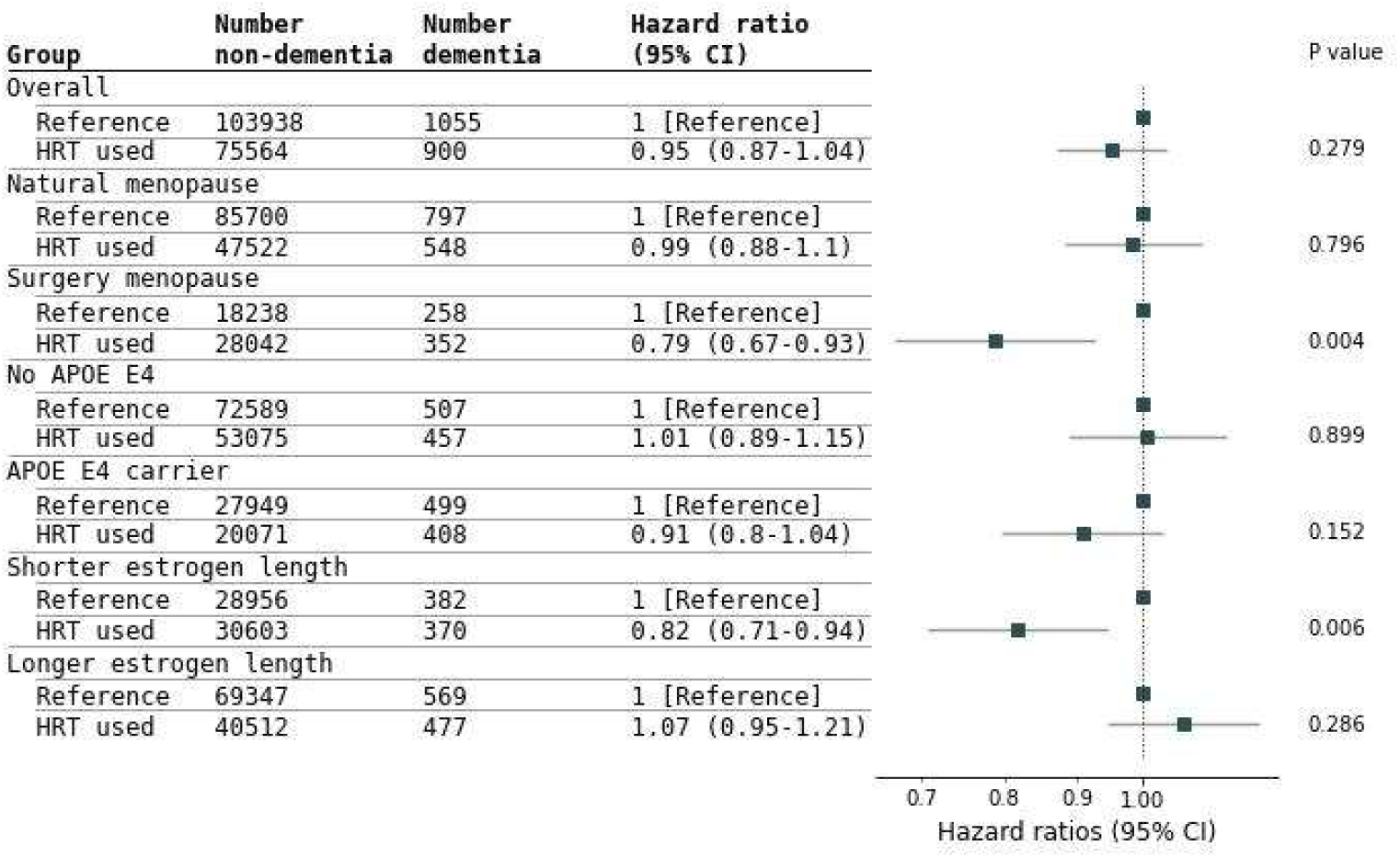
Risk of Incident non-AD Stratified by Groups. Abbreviations: APOE, apolipoprotein E. All results from a Cox proportional hazards regression adjusted for: age, education, smoking status, systolic blood pressure, ethnicity, BMI, cholesterol, Townsend deprivation index, diabetes, whether taking cholesterol lowering medication, whether taking anti-hypertensive medications. The reference for each group is those who never took HRT or took HRT but started and stopped at the same age. The HRT used women used HRT for at least one year or were still taking HRT at baseline. Natural menopause is women who reported cessation of periods and who had not have a bilateral oophorectomy or a hysterectomy. Surgical menopause are women who had either a bilateral oophorectomy or hysterectomy. No APOE ε4 women did not have any ε4 alleles. APOE ε4 carriers had either one or two ε4 alleles. Estrogen length is defined as time between menarche and the earliest of age at menopause, age at hysterectomy or age at bilateral oophorectomy. Shorter estrogen length is less than 35 years of estrogen length. Longer estrogen length is 35 years or more of estrogen length.

### Association between age at first use of HRT and dementia risk

The association between HRT and dementia risk was dependent on the age of starting HRT (Figure 4). For all post-menopausal women starting HRT between 46-50 years (inclusive) or 51-56 years (inclusive) was associated with reduced dementia risk with HRs of 0.87 (95% CI, 0.80-0.90, P=0.002) and 0.77 (95% CI, 0.69-0.86, P<0.001) respectively. For women who had a natural menopause starting HRT between 51-56 years was associated with reduced dementia risk with a HR of 0.82 (95% CI, 0.73-0.93, P=0.001) but starting HRT between 46-50 years was not significantly associated with reduced risk. Starting HRT either earlier than 47 years or later than 56 years was associated with a non-significant increased dementia risk. For women who had surgical menopause, HRT was associated with reduced dementia risk when started between 46-50 years and 51-56 years with HRs of 0.71 (95% CI, 0.61-0.82, P<0.001) and 0.61 (95% CI, 0.49-0.77, P<0.001) respectively.

**Figure 4.**
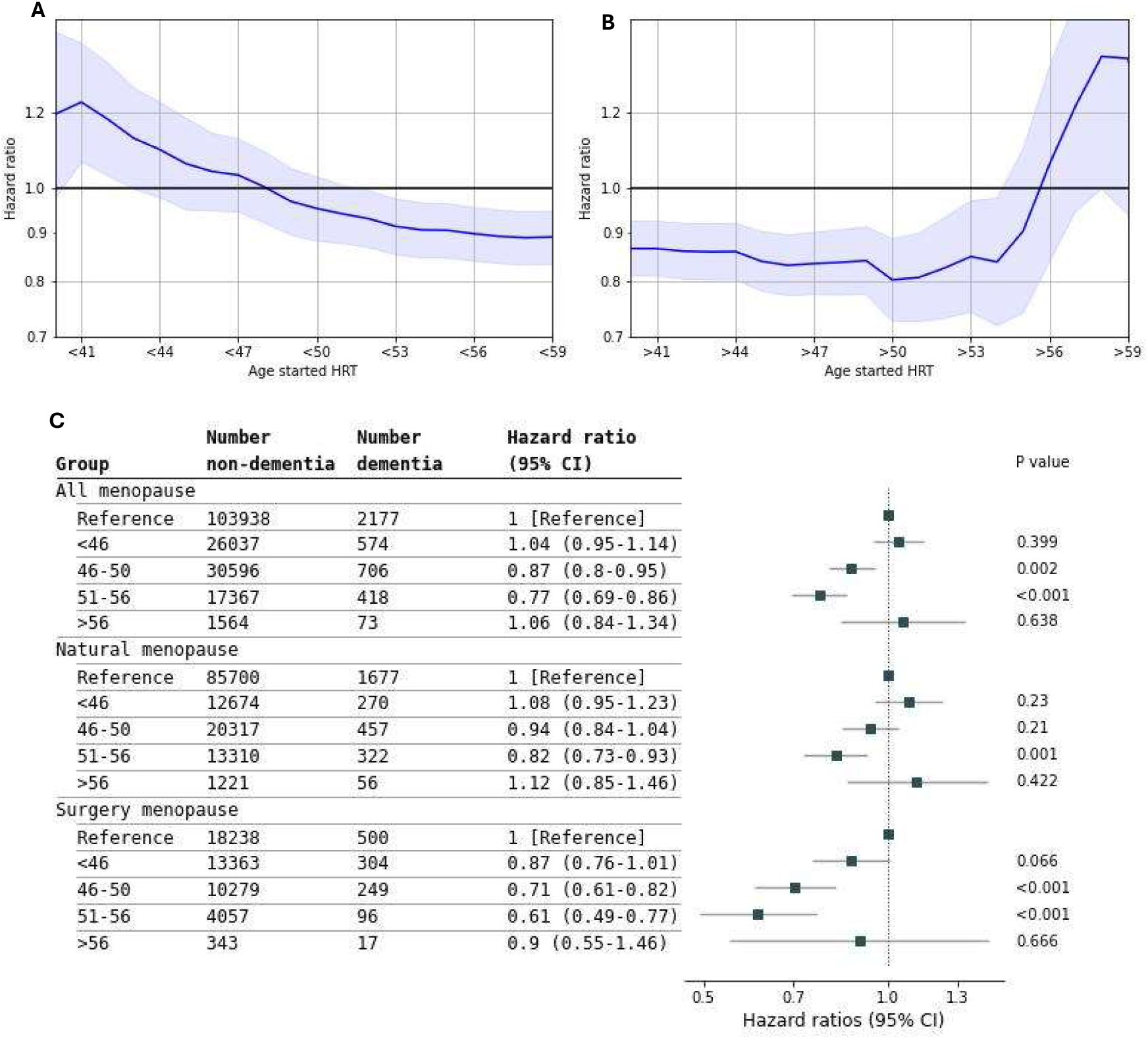
Risk of Incident Dementia by Starting Age of HRT. A) Hazard ratio for those that started using HRT before the stated age with the reference those women who never used HRT. Solid line is the hazard ratio and the shaded area is the 95% confidence intervals. B) Hazard ratio for those that started using HRT after the stated age with the reference those women who never used HRT. Solid line is the hazard ratio and the shaded area is the 95% confidence intervals. C) The starting age of HRT is split into four groups: those who started before 46 years, between 46-50 years (inclusive), between 51-56 years (inclusive) and those who started after 56 years. As well as all the women who had had menopause (All menopause) there are also stratifications into those who had a natural menopause or a surgical menopause (bilateral oophorectomy or hysterectomy).

### Sensitivity analyses

There were 9,065 women who used HRT for less than one year (starting age and age at last use are the same) who are included in the reference (non-HRT use) group. If these women are excluded from the analysis the results do not change significantly (eFigure 4 in the Supplement).

The proportion of women using HRT is higher in older women; in addition, younger women at the baseline assessment may start using HRT when they are older. Removing younger women from the analysis reduces these two effects and the overall HRT association remains similar (eFigure 5 in the Supplement).

Women who are diagnosed with dementia soon after baseline may have already had cognitive impairment which may affect the quality of the data provided; we removed women diagnosed with dementia within 2, 4, 6, and 8 years after baseline assessment (eFigure 6 in the Supplement) with no significant change in the HRT effect for any of these analyses.

The impact of surgery type and early versus late menarche and menopause are presented in eFigure 7, 8, 9 and 10 in the Supplement. We defined *APOE ε4* carriers as those with 1 or 2 ε4 alleles, in eFigure 11 of the Supplement we show hazard ratios for separated carriers of the *APOE ε2, ε3* and *ε4* alleles. In the main analysis *APOE ε2ε4* is included within the *APOE ε4* carrier group. As the *APOE ε2* allele is associated with reduced dementia risk, we reanalysed dementia risk with HRT with the *ε2ε4* carriers removed (eFigure 12 in the Supplement). There were no significant changes in results. We also introduced an interaction term into the Cox model between HRT and APOE ε4 status; with no significant associations (eFigure 13 in the Supplement).

The analysis of effect of age of starting HRT was repeated for AD and non-AD with results in eFigure 14 and 15 of the Supplement. Results for starting HRT stratified by women who had a natural menopause and surgical menopause are in eFigures 16 and 17 of the Supplement.

The main analyses were repeated with two different sets of covariates: no covariates; and only age, education and smoking status as covariates. These results are reported in eFigures 18-25 in the Supplement. The pattern of the results was similar to the fully adjusted models used in the primary analyses.

## Discussion

This UK Biobank analysis is the most comprehensive prospective analysis to date of the association between HRT and incident dementia and its potential mediators. We showed that HRT was associated with a 10% reduced dementia risk, and modulated by *APOE* genotype, menopause ‘type’ and lifetime exposure to natural estrogen. Compared to non-users a 32% reduced risk was evident in *APOE ε4* women who had undergone surgical menopause and had a short life-time estrogen, exposure. Significant associations between HRT and dementia were only evident in the women who had initiated HRT between 46-56 years supporting the ‘window of opportunity’ hypothesis for HRT benefits.

Early observation studies reported that estrogen therapy was associated with reduced dementia^36^. However, the only RCT with incident dementia conducted to date, the WHIMS, observed an increased incidence of dementia over 5 years following estrogen plus progestogen HRT in participants >65 years^10^. Subsequent RCTS in younger participants, but nevertheless mainly in early post-menopause, have observed little effect of HRT on cognitive outcomes^37,38^. Our finding provide nuance to the importance of HRT timing, supporting the healthy cell bias hypothesis^39^, of greater benefit in the absence of significant existing neuropathology if HRT is taken between 46-56 years which typically represent the perimenopausal and early postmenopausal periods. Mechanistic analyses suggest that the shorter the delay between menopause and onset of treatment, the higher chance of favourable effects of HRT on the brain^2,40^. HRT timing has also been identified as important for cardiovascular benefit.^41^

The basis for our observation of stronger associations and potential benefit of HRT in surgical versus natural menopause needs consideration as it may in part reflect the HRT formulation. In natural menopause a combined estrogen-progestogen HRT is now typically prescribed with progestogen preventing endothelial hyperplasia in women with a functional uterus. In surgical menopause a hysterectomy is common and estrogen only HRT formulations subsequently prescribed. The WHIMS and large cohort studies have consistently reported that estrogen only therapy is associated with lower dementia risk compared to the combined HRT^9,10,42^. Lack of information on HRT formulation is a limitation of the current analysis.

In both menopausal groups we observed the strongest associations in carriers of the *APOE ε4* allele which is consistent with our previously smaller scale cross-section analysis in the EPAD cohort where the association between HRT and cognition and brain volumes in the media temporal lobe were only evident in *APOE ε4*^43^. The *APOE ε4* allele is associated with accelerated neurocognitive decline and higher dementia risk, with an allele frequency of 20-25% in the general population versus 50-75% in AD^44^. Evidence of greater benefit in *APOE ε4* is potentially due to the overlapping molecular targets of APOE and estrogen, with the *APOE ε4* allele negatively impacting synaptic plasticity, BBB function, glucose hypometabolism and neuroinflammation, relative to the wildtype genotype^45^.

### Strengths and limitations

A strength of our study is that the amount of available data, alongside the follow-up for dementia diagnosis, gives us a large dataset to assess the relationship between HRT and dementia risk, with almost 4000 incident cases over up to 16 years follow up. We were able to stratify women into various subgroups to show that the association between HRT and dementia risk varies considerably. The availability of a significant amount of additional data in UK Biobank allowed a comprehensive sensitivity analysis to multiple potential factors and confounders.

A limitation of our study is that results are associations and not causal. It may be that factors other than HRT are involved that result in both HRT and reduced dementia risk. As mentioned above the lack of information on HRT formulation (estrogen only or estrogen plus progestogen), dose and mode of administration (topical, oral, vaginal) in UK Biobank prevents any analysis on the impact of these important variables. UK Biobank is also based mostly on women of European descent and tends to be drawn from some biased social classes^46^. In addition, although UK Biobank is a large study we lack statistical power to draw strong conclusions when we stratify by groups or dementia sub-types. Diagnoses are primarily from secondary care (hospital inpatient admissions); future work integrating more primary care diagnoses would strengthen the analysis.

## Conclusions

In UK Biobank participants, HRT was associated with reduced dementia risk with larger associations for AD than non-AD. The associations are greatest in those who had undergone surgical menopause, and when HRT is initiated between ages 46-56 years. For women who had a natural menopause, HRT associations were only evident in *APOE ε4* carriers with a low lifetime exposure to estrogen. The findings support a more personalised approach to HRT prescribing for reducing population dementia risk.

## Data Availability

UK Biobank data is available to researchers through application to: https://www.ukbiobank.ac.uk/.

## Acknowledgements

This work was funded by a UK Biotechnology and Biological Sciences Research Council (BBSRC/UKRI) project grant, BB/X002209/1. JLA is funded by an NIHR Advanced Fellowship (NIHR301844). JR is funded by Alzheimer’s Research UK. Alzheimer’s Research UK (DJL), the National Institute for Health and Care Research (NIHR) Applied Research Collaboration South West Peninsula (DJL), and the NIHR Exeter Biomedical Research Centre (DJL).

This research has been conducted using the UK Biobank Resource under Application Number 14631. This work uses data provided by patients and collected by the NHS as part of their care and support

