## Supplementary Material for "Hormone replacement therapy and dementia risk among postmenopausal women: evidence from the UK Biobank"

**eMethods**

Included participants

Figure 1 Flowchart of included participants in the overall analysis

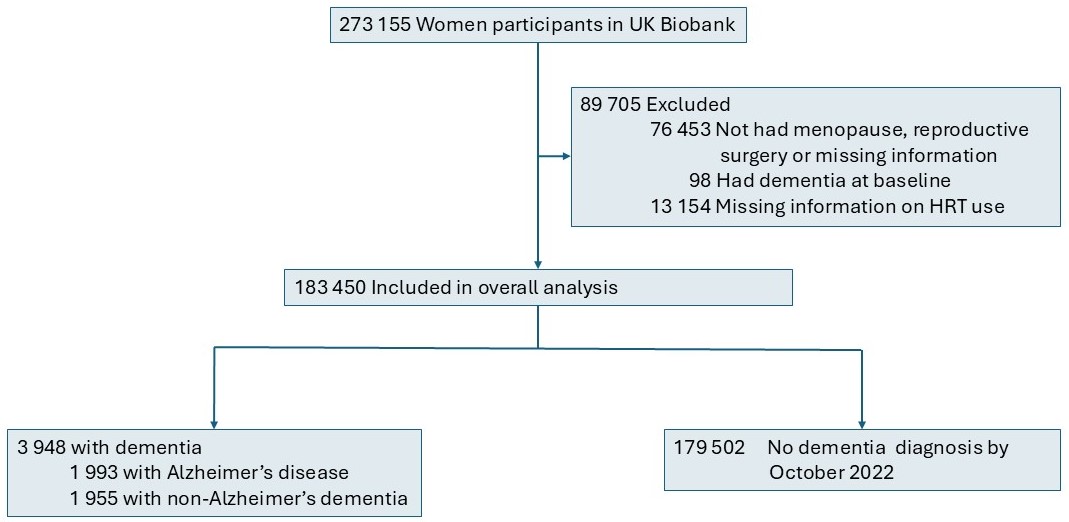

Study design and participants

UK Biobank has ethical approval from the North West Multi-centre Research Ethics Committee (reference 11/NW/0382) and all participants provided informed consent through electronic signature at baseline assessment. Because data were deidentified, this study did not require additional approval. Access to UK Biobank was granted under Application Number 14631.

Incident dementia

We ascertained dementia diagnoses from three sources of linked medical records data: 1) hospital inpatient records containing data on primary and secondary diagnoses obtained from the Hospital Episode Statistics (HES) for England, Scottish Morbidity Record data for Scotland, and the Patient Episode Database for Wales; 2) death register data on underlying/contributory cause of death provided by the National Health Service Digital for England and Wales and the Information and Statistics Division for Scotland; and 3) primary care (General Practice, GP) diagnosis data were available in 45% of the cohort, up to 2016 or 2017 depending on data provider (<https://biobank.ndph.ox.ac.uk/ukb/label.cgi?id=3000>). HES and death record diagnoses were recorded using the *International Classification of Diseases* (*ICD*) coding system. GP data are recorded as Read2 or CTV3 codes, depending on the data provider.

HRT definition

The exposure of interest was use of HRT as defined by UKBB fields: ever used HRT (UKBB ID 2814), age started HRT (UKBB ID 3536) and age last used HRT (UKBB ID 3546). We define no HRT use as those women saying they never used HRT or saying they did use HRT but then stating the start and last use age as the same (i.e. started and stopped HRT at the same age). We excluded from all analysis women who did not know if they used HRT or refused to answer, and also if they said they did use HRT but did not answer the age they started or last used HRT.

Additional stratifications by APOE4 status, age of menarche, age at menopause, length of oestrogen and reproductive surgery

We stratified the results by APOE ɛ4 status: we defined a APOE ɛ4 carrier as having one or two APOE ɛ4 alleles. APOE alleles are denoted as 0 or 1 with 0: APOE ɛ3/3, APOE ɛ2/2, APOE ɛ2/3 and 1: APOE ɛ3/4, APOE ɛ2/4, APOE ɛ4/4. We also performed a sensitivity analysis removing the APOE ɛ2/4 carriers entirely.

We additionally stratified by age at menarche (UKBB ID 2714), age at menopause and years between menarche and menopause. Age of spontaneous (natural) menopause is given by UKBB ID 3581. For women who had either a hysterectomy and/or bilateral oophorectomy (given by UKBB IDs 2724, 3591, 2834) we defined the age of menopause as the earliest age of surgery or stated menopause age (3581). We stratified into pairs of groups for age of menarche (14 or younger, 15 or older), age of menopause (44 or younger, 45 or older) and length of natural estrogen (34 years or fewer, 35 years or more). Any that have missing data on age of menarche or age of menopause were removed from that stratification.

We also stratified by age of starting HRT to investigate a possible window of opportunity. The reference class is the same as for the other analyses: women who never took HRT or started and stopped at the same age. We compare the reference class with women who started HRT between the ages of 46-50 years (inclusive) and (separately) 51-56 years (inclusive). We also explore the hazard ratio changes if we compare all women who started HRT before a set if ages between 41 and 59 years. Separately we do the same but for all women who started HRT after that age. For these analyses we additionally stratified into women who had any-menopause, a natural menopause, or a surgery menopause.

In Table 1 are the covariates used in the adjusted model, an additional column specifies which variables were used in the partially adjusted model

Table 1 Covariates used in the models

| Covariate | Model | ID | Details |
| --- | --- | --- | --- |
| Age | 2,3 | 21022 | Age at baseline |
| Education | 2,3 | 6138 | Five categories: none of the above; CSEs or equivalent & O levels/GCSEs or equivalent; A levels/AS levels or equivalent; NVQ or HND or HNC or equivalent; College or University degree & Other professional qualification. |
| Smoking | 2,3 | 20116 | Never smoked=0, smoked ever/current=1 |
| Systolic blood pressure | 3 | 4080 | No alterations |
| Ethnicity | 3 | 21000 | Six categories: white; black; Asian; mixed; Chinese; other |
| BMI | 3 | 21001 | No alterations |
| Cholesterol | 3 | 26037 | No alterations |
| Townsend | 3 | 22189 | No alterations |
| Diabetes | 3 | 2443 | No alterations |
| Cholesterol lowering medication | 3 | 6153 | No alterations |
| Anti-hypertensive medication | 3 | 6153 | No alterations |

**Supplementary Results**

Combinations of surgery, length of natural estrogen and APOE status

Figure 2 A decision tree for the hazard ratios given for combinations of different stratifications for use of HRT on dementia.

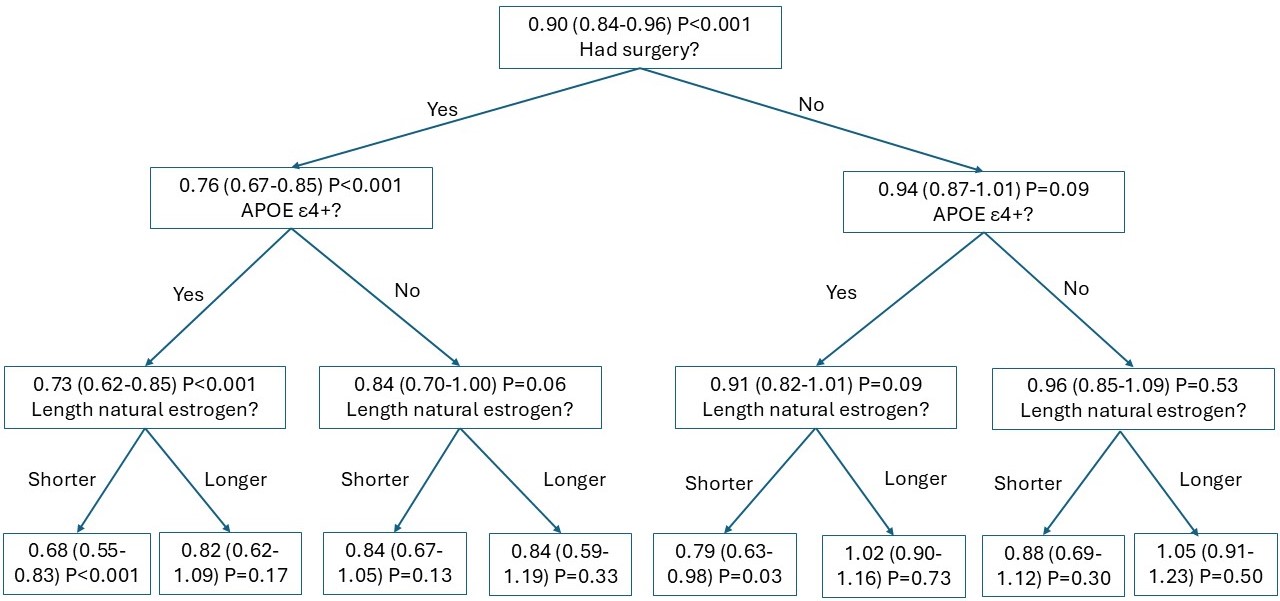

The number in each box is the hazard ratio for that stratification. The question in each box leads to the next (additional) stratification. The top of the tree is overall use of HRT, the first split is shown for surgery, the second for APOE ɛ4 status and the third for length of estrogen.

Figure 3 Decisions trees for menopause type, length of estrogen and APOE ɛ4 status.

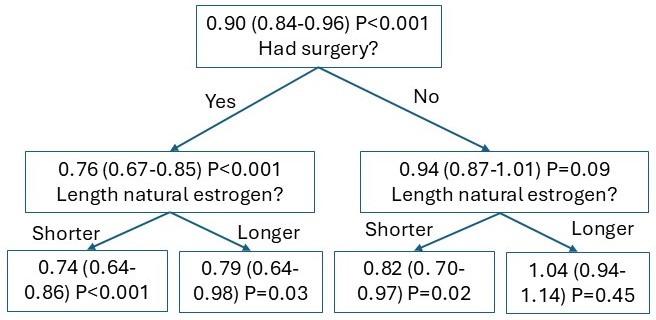

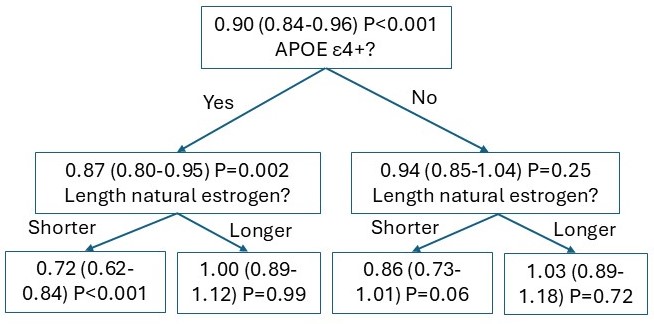

These decision trees show different ordering of the stratifications.

Different definitions of HRT use

Figure 4 Different definition of HRT use

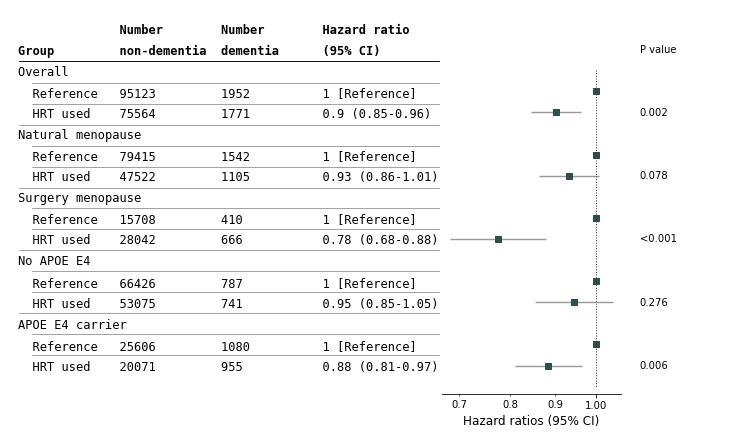

Different definition of HRT use for the adjusted model and all dementia. Those women who used HRT but started and stopped at the same age were excluded from the analysis instead of being including in no HRT use.

Removing younger women

Figure 5 Removing women younger than the specified age for the adjusted model and all dementia

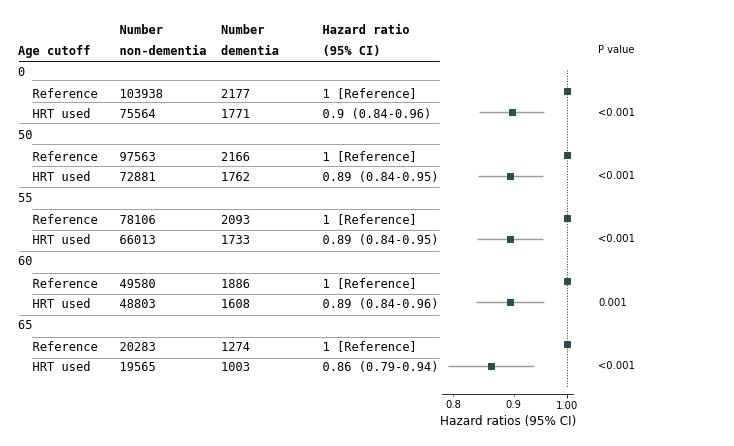

For each group labelled by 0, 50, 55, 60, 65, all women under that age were excluded from the analysis. For 0 all women were included. For 50 any women under 50 years old were excluded from the analysis. Results shown are for dementia and all menopause status.

Removing women diagnosed near baseline

Figure 6 Removing women diagnosed with dementia within stated number of years of baseline for the adjusted model and all dementia.

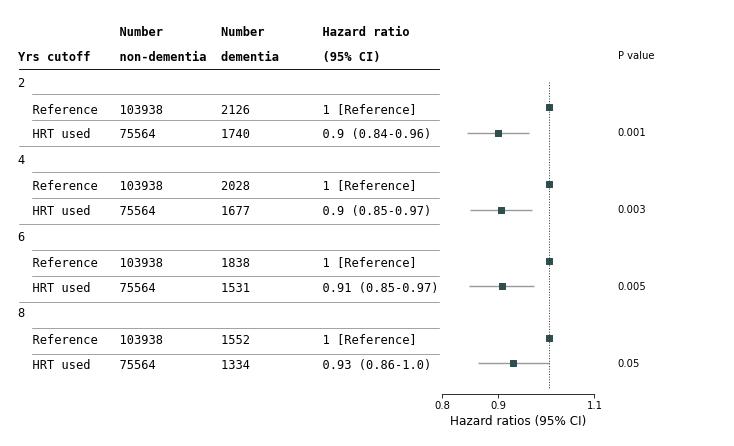

The cutoffs are set so that any women diagnosed within less than that number of years from their baseline assessment was excluded from that group. For example at 2 yrs cutoff all women who were diagnosed less than 2 years after their baseline assessment were excluded from the analysis.

Surgery subtypes

Figure 7 Surgery subtypes for dementia and the adjusted model

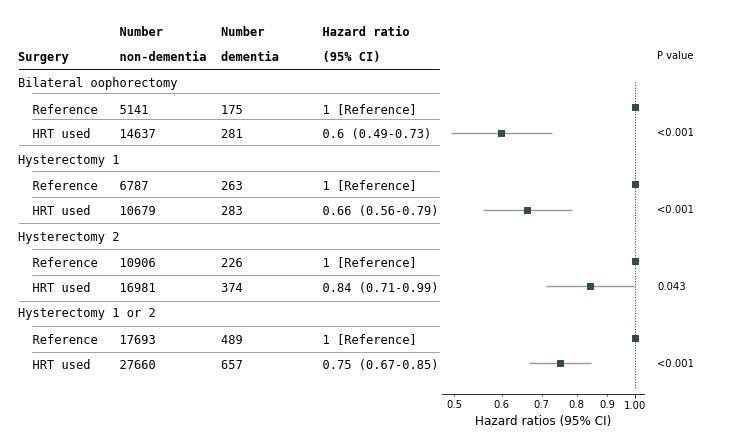

Hysterectomy 1 are women who answered the question “Have you had a hysterectomy (womb removed)?” (UK Biobank ID 3591). Hysterectomy 2 are women who, when asked “Have you had your menopause (periods stopped)?” (UK Biobank ID 2724) answered: “Not sure - had a hysterectomy”

Stratification by early/late menarche and early/late menopause

Figure 8 Risk of Incident Dementia Stratified by Early/Late Menarche/Menopause for all-menopause

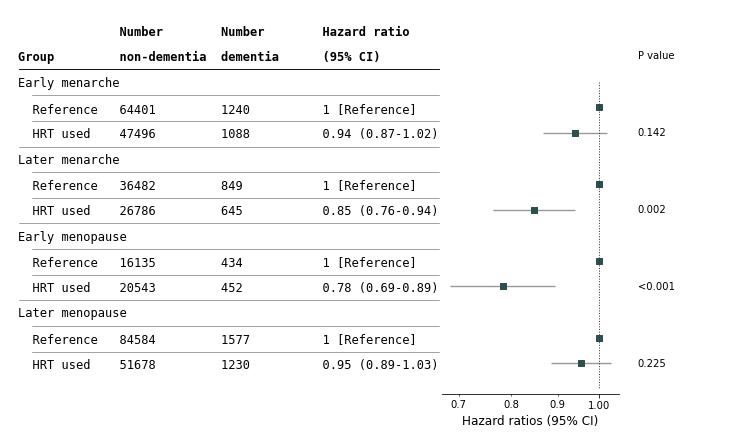

Early menarche is defined as under 14 and later menarche 14 or older. Early menopause is defined as under 45 and later menopause as 45 or older.

Figure 9 Risk of Incident Dementia Stratified by Early/Late Menarche/Menopause for natural-menopause

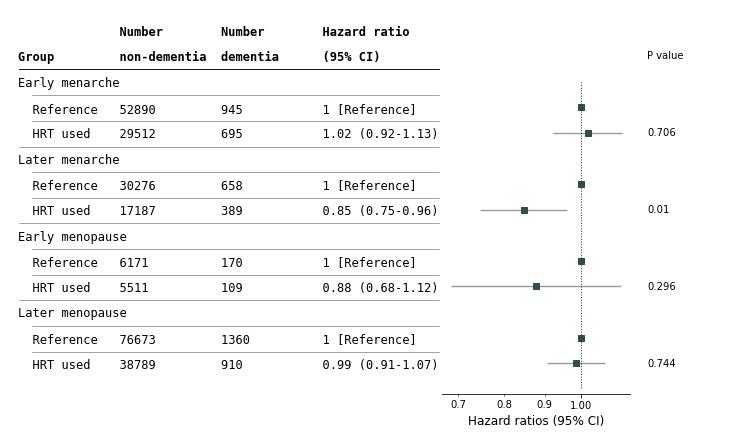

Early menarche is defined as under 14 and later menarche 14 or older. Early menopause is defined as under 45 and later menopause as 45 or older.

Figure 10 Risk of Incident Dementia Stratified by Early/Late Menarche/Menopause for surgery-menopause

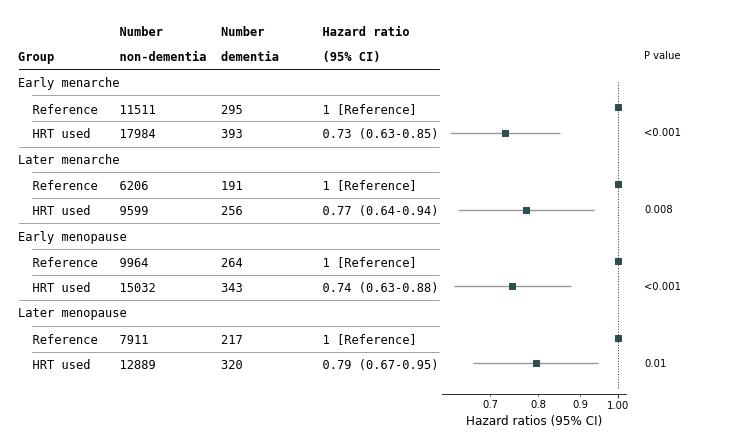
 Early menarche is defined as under 14 and later menarche 14 or older. Early menopause is defined as under 45 and later menopause as 45 or older.

Different APOE alleles

Figure 11 Hazard ratios for different combinations of APOE for the adjusted model and all dementia

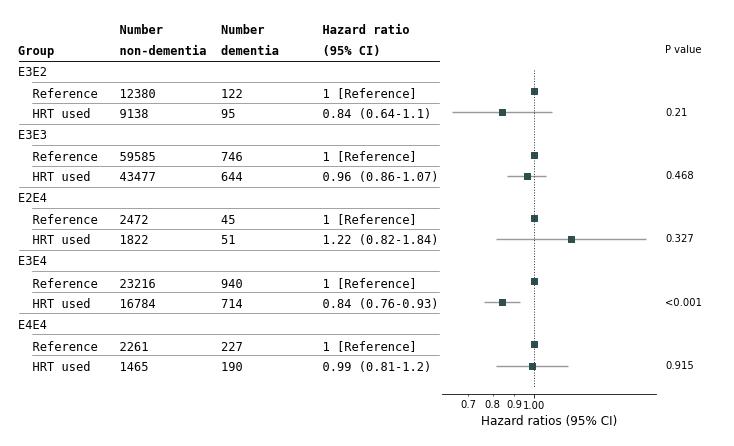

APOE E2E2 are excluded as there are too few to produce a model.

Excluding APOE ɛ2ɛ4 carriers

Figure 12 Risk of Incident Dementia Stratified by Groups excluding APOE ɛ2ɛ4 carriers

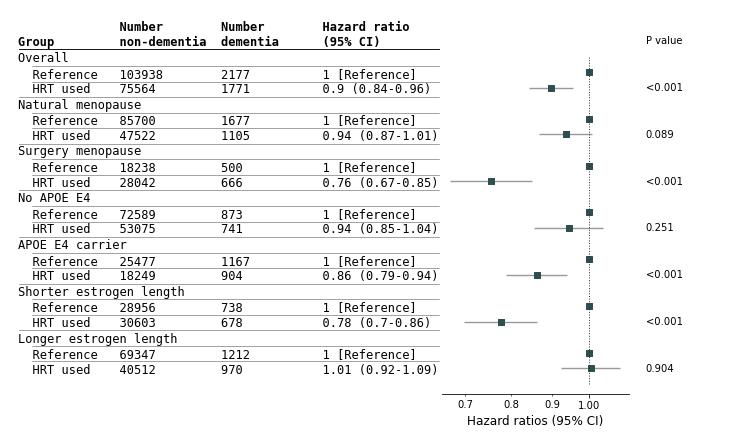

APOE and HRT interactions

Figure 13 HRT-APOE interaction terms for model3 and all dementia

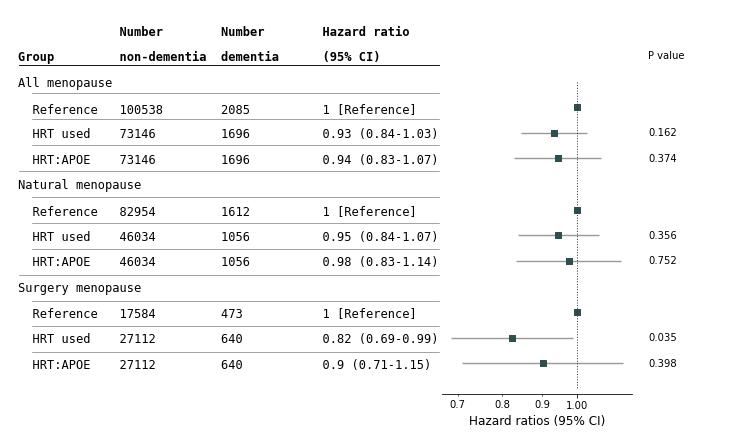

HRT:APOE is the interactive term.

HRT start age for AD and non-AD

Figure 14 Risk of Incident Alzheimer’s disease by Starting Age of HRT

**b**

**a**
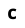

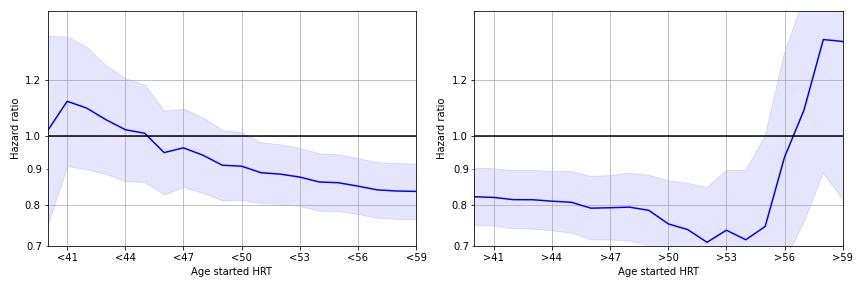

**c**

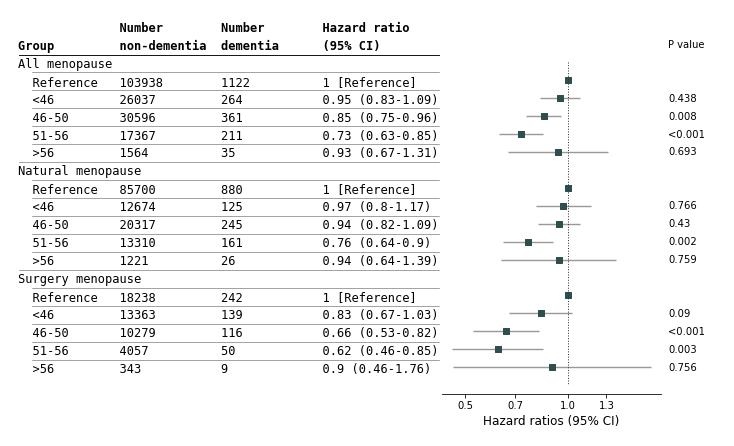

A) Hazard ratio for those that started using HRT before the stated age with the reference those women who never used HRT or used HRT for less than one year. Solid line is the hazard ratio and the shaded area is the 95% confidence intervals. B) Hazard ratio for those that started using HRT after the stated age with the reference those women who never used HRT or used HRT for less than one year. Solid line is the hazard ratio and the shaded area is the 95% confidence intervals. C) The starting age of HRT is split into four groups: those who started before 46, between 46-50 (inclusive), between 51-56 (inclusive) and those who started after 56. As well as all the women who had had menopause (All menopause) there are also stratifications into those who had a natural menopause (no bilateral oophorectomy or hysterectomy) and a surgery menopause (bilateral oophorectomy or hysterectomy).

Figure 15 Risk of Incident non-Alzheimer’s dementia by Starting Age of HRT

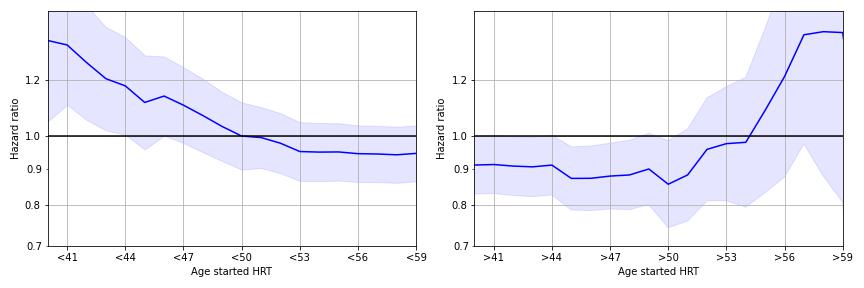

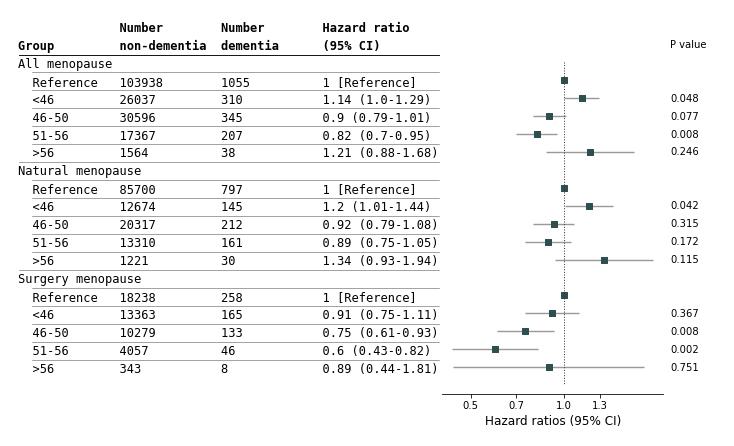

A) Hazard ratio for those that started using HRT before the stated age with the reference those women who never used HRT or used HRT for less than one year. Solid line is the hazard ratio and the shaded area is the 95% confidence intervals. B) Hazard ratio for those that started using HRT after the stated age with the reference those women who never used HRT or used HRT for less than one year. Solid line is the hazard ratio and the shaded area is the 95% confidence intervals. C) The starting age of HRT is split into four groups: those who started before 46, between 46-50 (inclusive), between 51-56 (inclusive) and those who started after 56. As well as all the women who had had menopause (All menopause) there are also stratifications into those who had a natural menopause (no bilateral oophorectomy or hysterectomy) and a surgery menopause (bilateral oophorectomy or hysterectomy).

Window of opportunity for natural and surgical menopause

Figure 16 Risk of Incident Dementia by Starting Age of HRT for women who had a natural menopause

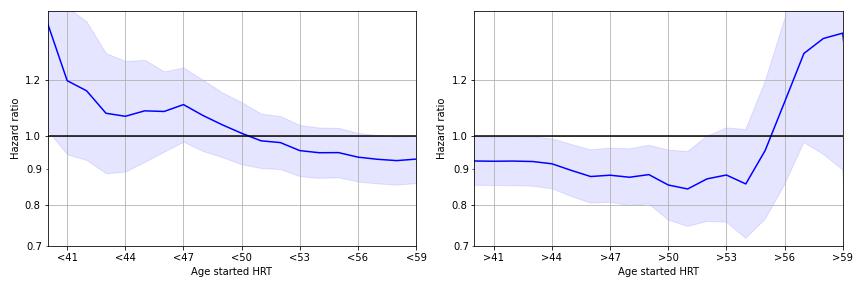

Stratified by women who had a natural menopause.

Figure 17 Risk of Incident Dementia by Starting Age of HRT for women who had a surgery menopause

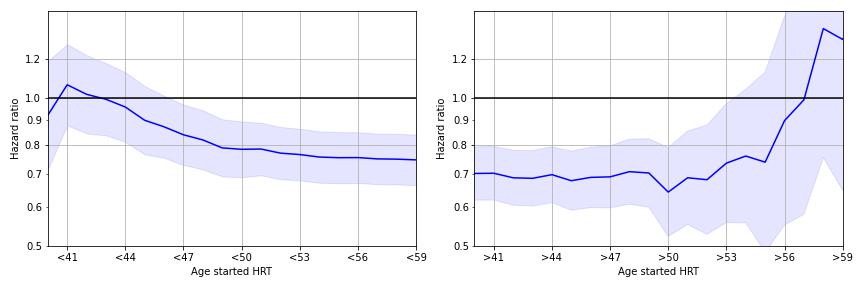

Stratified by women who had a surgery menopause. The y-axis scale is different than other equivalent plots.

Recreation of main paper plots with no covariates

Figure 18 Risk of Incident Dementia Stratified by Groups

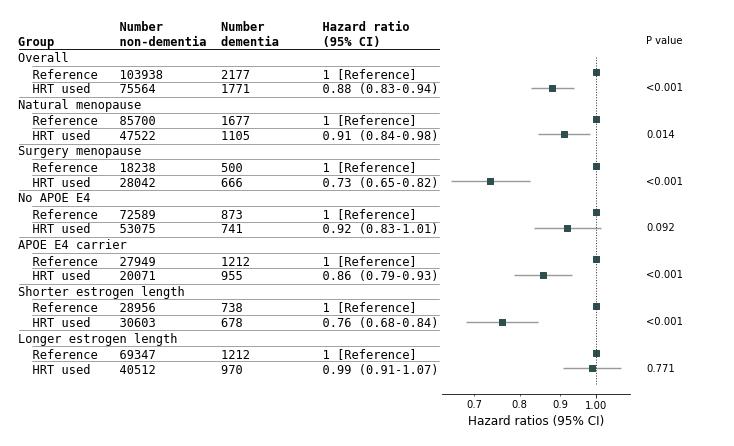

Abbreviations: APOE, apolipoprotein E. All results from a Cox proportional hazards regression not additionally adjusted. The reference for each group is those who never took HRT or took HRT but started and stopped at the same age. The women defined as using HRT had used HRT for at least one year or were still taking HRT at baseline. Natural menopause is women who reported cessation of periods and who had not had a bilateral oophorectomy or a hysterectomy. Surgery menopause are women who had either a bilateral oophorectomy or hysterectomy. No APOE ɛ4 women did not have any ɛ4 alleles. APOE ɛ4 carriers had either one or two ɛ4 alleles. Estrogen length is defined as time between menarche and the earliest of age at menopause, age at hysterectomy or age at bilateral oophorectomy. Shorter estrogen length is less than 35 years of natural estrogen length. Longer estrogen length is 35 years or more of natural estrogen length.

Figure 19 Risk of Incident AD Stratified by Groups

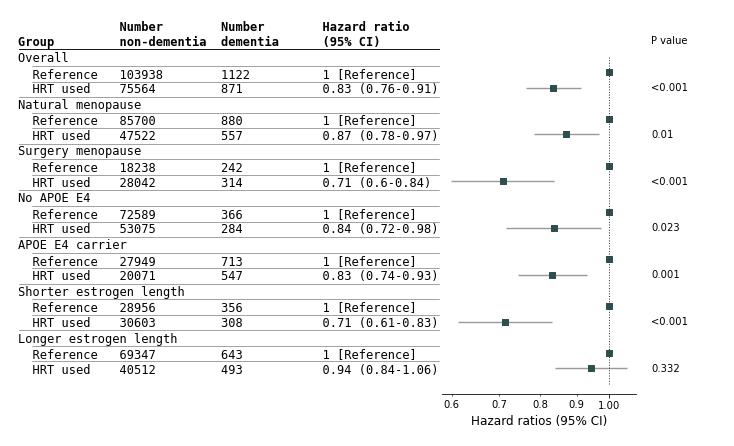

Abbreviations: APOE, apolipoprotein E. All results from a Cox proportional hazards regression not additionally adjusted. The reference for each group is those who never took HRT or took HRT but started and stopped at the same age. The HRT used women used HRT for at least one year or were still taking HRT at baseline. Natural menopause is women who reported cessation of periods and who had not have a bilateral oophorectomy or a hysterectomy. Surgery menopause are women who had either a bilateral oophorectomy or hysterectomy. No APOE ɛ4 women did not have any ɛ4 alleles. APOE ɛ4 carriers had either one or two ɛ4 alleles. Estrogen length is defined as time between menarche and the earliest of age at menopause, age at hysterectomy or age at bilateral oophorectomy. Shorter estrogen length is less than 35 years of estrogen length. Longer estrogen length is 35 years or more of estrogen length.

Figure 20 Risk of Incident non-AD Stratified by Groups

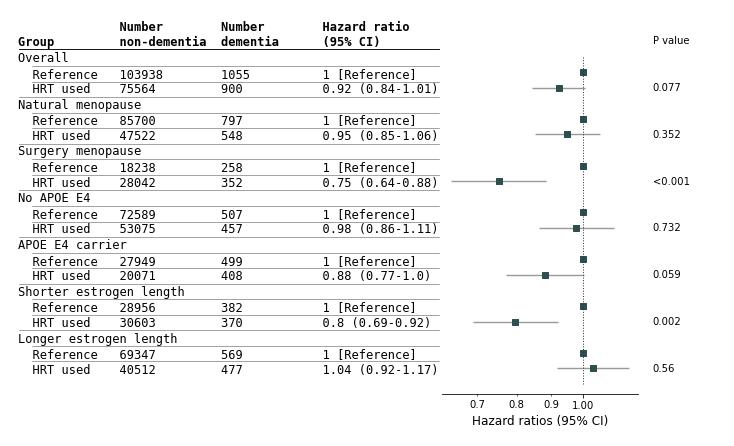

Abbreviations: APOE, apolipoprotein E. All results from a Cox proportional hazards regression not additionally adjusted. The reference for each group is those who never took HRT or took HRT but started and stopped at the same age. The HRT used women used HRT for at least one year or were still taking HRT at baseline. Natural menopause is women who reported cessation of periods and who had not have a bilateral oophorectomy or a hysterectomy. Surgery menopause are women who had either a bilateral oophorectomy or hysterectomy. No APOE ɛ4 women did not have any ɛ4 alleles. APOE ɛ4 carriers had either one or two ɛ4 alleles. Estrogen length is defined as time between menarche and the earliest of age at menopause, age at hysterectomy or age at bilateral oophorectomy. Shorter estrogen length is less than 35 years of estrogen length. Longer estrogen length is 35 years or more of estrogen length.

Figure 21 Risk of Incident Dementia by Starting Age of HRT

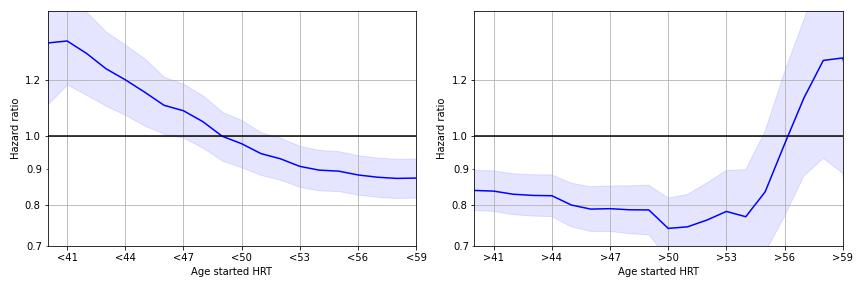

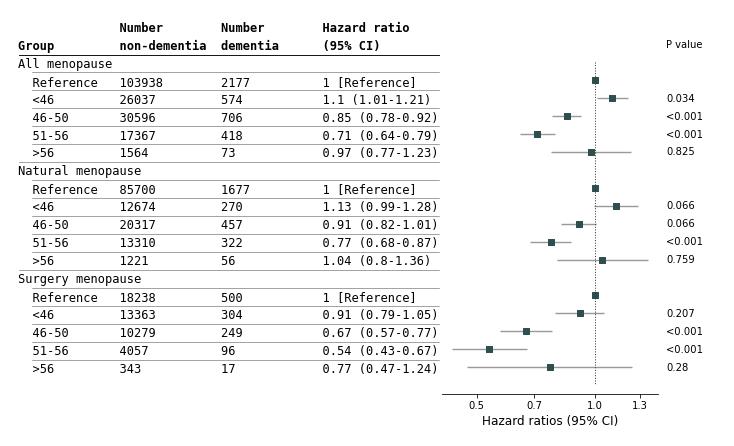

A) Hazard ratio for those that started using HRT before the stated age with the reference those women who never used HRT or used HRT for less than one year. Solid line is the hazard ratio and the shaded area is the 95% confidence intervals. B) Hazard ratio for those that started using HRT after the stated age with the reference those women who never used HRT or used HRT for less than one year. Solid line is the hazard ratio and the shaded area is the 95% confidence intervals. C) The starting age of HRT is split into four groups: those who started before 46, between 46-50 (inclusive), between 51-56 (inclusive) and those who started after 56. As well as all the women who had had menopause (All menopause) there are also stratifications into those who had a natural menopause (no bilateral oophorectomy or hysterectomy) and a surgery menopause (bilateral oophorectomy or hysterectomy).

Recreation of main paper plots with age, education and smoking status as covariates

Figure 22 Risk of Incident Dementia Stratified by Groups

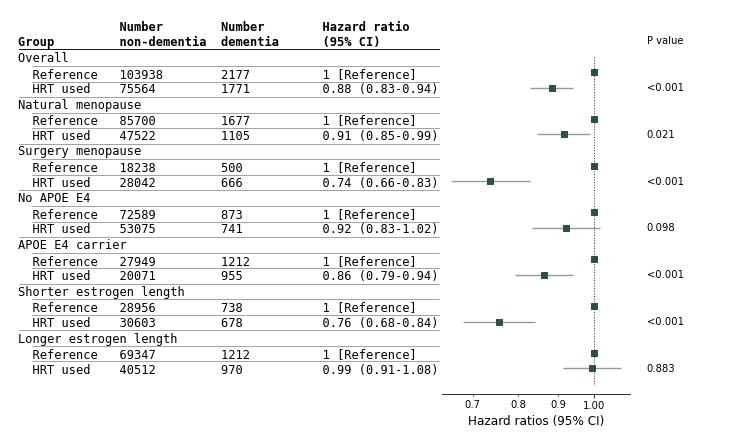

Abbreviations: APOE, apolipoprotein E. All results from a Cox proportional hazards regression adjusted for: age, education and smoking status. The reference for each group is those who never took HRT or took HRT but started and stopped at the same age. The women defined as using HRT had used HRT for at least one year or were still taking HRT at baseline. Natural menopause is women who reported cessation of periods and who had not had a bilateral oophorectomy or a hysterectomy. Surgery menopause are women who had either a bilateral oophorectomy or hysterectomy. No APOE ɛ4 women did not have any ɛ4 alleles. APOE ɛ4 carriers had either one or two ɛ4 alleles. Estrogen length is defined as time between menarche and the earliest of age at menopause, age at hysterectomy or age at bilateral oophorectomy. Shorter estrogen length is less than 35 years of natural estrogen length. Longer estrogen length is 35 years or more of natural estrogen length.

Figure 23 Risk of Incident AD Stratified by Groups

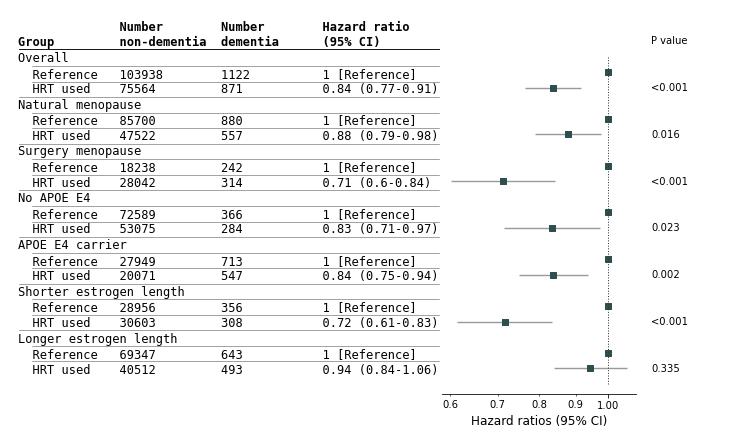

Abbreviations: APOE, apolipoprotein E. All results from a Cox proportional hazards regression adjusted for: age, education and smoking status. The reference for each group is those who never took HRT or took HRT but started and stopped at the same age. The HRT used women used HRT for at least one year or were still taking HRT at baseline. Natural menopause is women who reported cessation of periods and who had not have a bilateral oophorectomy or a hysterectomy. Surgery menopause are women who had either a bilateral oophorectomy or hysterectomy. No APOE ɛ4 women did not have any ɛ4 alleles. APOE ɛ4 carriers had either one or two ɛ4 alleles. Estrogen length is defined as time between menarche and the earliest of age at menopause, age at hysterectomy or age at bilateral oophorectomy. Shorter estrogen length is less than 35 years of estrogen length. Longer estrogen length is 35 years or more of estrogen length.

Figure 24 Risk of Incident non-AD Stratified by Groups

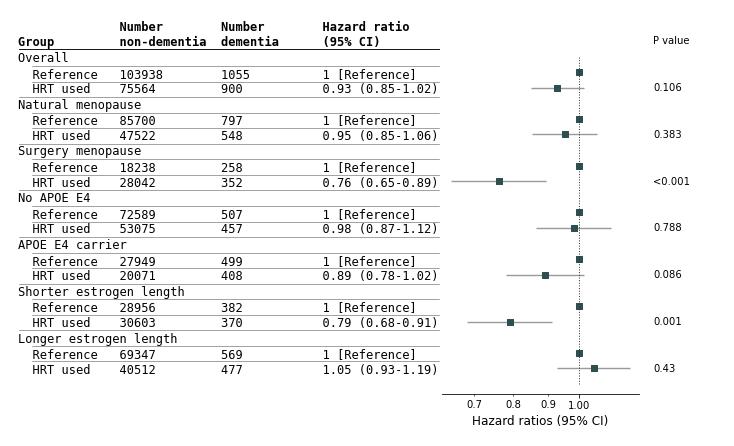

Abbreviations: APOE, apolipoprotein E. All results from a Cox proportional hazards regression adjusted for: age, education and smoking status. The reference for each group is those who never took HRT or took HRT but started and stopped at the same age. The HRT used women used HRT for at least one year or were still taking HRT at baseline. Natural menopause is women who reported cessation of periods and who had not have a bilateral oophorectomy or a hysterectomy. Surgery menopause are women who had either a bilateral oophorectomy or hysterectomy. No APOE ɛ4 women did not have any ɛ4 alleles. APOE ɛ4 carriers had either one or two ɛ4 alleles. Estrogen length is defined as time between menarche and the earliest of age at menopause, age at hysterectomy or age at bilateral oophorectomy. Shorter estrogen length is less than 35 years of estrogen length. Longer estrogen length is 35 years or more of estrogen length

Figure 25 Risk of Incident Dementia by Starting Age of HRT

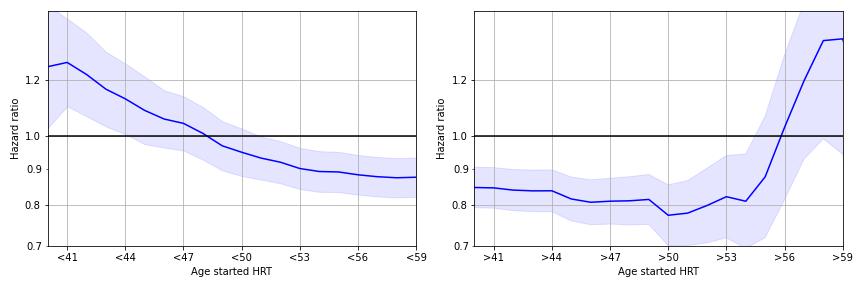

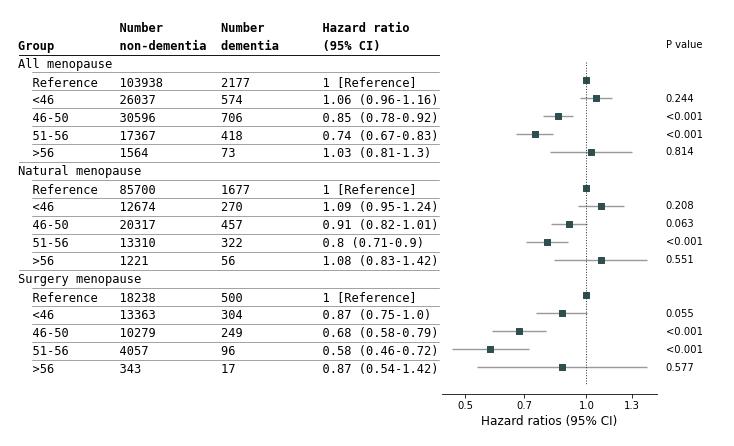

A) Hazard ratio for those that started using HRT before the stated age with the reference those women who never used HRT or used HRT for less than one year. Solid line is the hazard ratio and the shaded area is the 95% confidence intervals. B) Hazard ratio for those that started using HRT after the stated age with the reference those women who never used HRT or used HRT for less than one year. Solid line is the hazard ratio and the shaded area is the 95% confidence intervals. C) The starting age of HRT is split into four groups: those who started before 46, between 46-50 (inclusive), between 51-56 (inclusive) and those who started after 56. As well as all the women who had had menopause (All menopause) there are also stratifications into those who had a natural menopause (no bilateral oophorectomy or hysterectomy) and a surgery menopause (bilateral oophorectomy or hysterectomy).
